# Macro-Level Drivers of SARS-CoV-2 Transmission: A Data-Driven Analysis of Factors Contributing to Epidemic Growth During the First Wave of outbreaks in the United States

**DOI:** 10.1101/2021.06.23.21259394

**Authors:** Matthew J Watts

## Abstract

**Background:** Many questions remain unanswered about how SARS-CoV-2 transmission is influenced by aspects of the economy, environment, and health. A better understanding of how these factors interact can help us to design early health prevention and control strategies, and develop better predictive models for public health risk management of SARS-CoV-2. This study examines the associations between COVID-19 epidemic growth and macro-level determinants of transmission such as climate, socio-economic factors, demographic factors, and population health, during the first wave of outbreaks in the United States.

**Methods:** A spatial-temporal data-set was created by collating information from a variety of data sources including the Johns Hopkins University’s Centre for Systems Science and Engineering, the United States Census Bureau, the USDA Economic Research Service, the United States EPA, the National Climatic Data Center, the CDC and the Oxford COVID-19 Government Response Tracker (OxCGRT). A unique data-driven study design was implemented that allows us to assess the relationship between COVID-19 case and death epidemic doubling times and explanatory variables using a Generalized Additive Model (GAM).

**Results:** The main factors associated with case doubling times are higher population density, home overcrowding, manufacturing, and recreation industries. Poverty was also an important predictor of faster epidemic growth perhaps because of factors associated with in-work poverty-related conditions, although poverty is also a predictor of poor population health which is likely driving case and death reporting. Air pollution and diabetes were other important drivers of case reporting. Warmer temperatures are associated with slower epidemic growth, which is most likely explained by human behaviors associated with warmer locations i.e ventilating homes and workplaces. and socializing outdoors. The main factors associated with death doubling times were population density, poverty older age, diabetes, and air pollution. Temperature was also slightly significant slowing death doubling times.

**Conclusions:** Such findings help underpin current understanding of the disease epidemiology and also supports current policy and advice recommending ventilation of homes, work-spaces, and schools, along with social distancing and mask-wearing. The results also suggest that states which adopted more stringent containment measures early on did have some success suppressing the virus. We can presume that if this was replicated at a federal level, much better outcomes would have been observed across the United States.

## 1 Introduction

The current COVID-19 pandemic is posing severe challenges to health systems, societies, and economies worldwide. At the time of writing, the The SARS-CoV-2 virus has already infected more than 175 million people globally and caused 3.7 M deaths. In addition, the long-term health impacts on those who have recovered from the SARS-CoV-2 infection are still unknown [1]. Approximately a sixth of the total deaths - more than 600,000 - occurred in the United States (US), the country that currently stands with the highest number of fatalities.

In the US and in other European countries like the the United Kingdom, governments, and public health systems were initially caught off guard by the sudden and rapid spread of the virus. This was partly due to a lack of political preparedness and a coherent strategy, lack of public health resources after years of cuts to public health budgets, or to the adoption of the wrong or no policy in terms of mask-wearing, contact tracing, border controls, or lack of testing to detect community transmission [2–7]. Furthermore, the scientific community took some time to reach a general consensus about the modes of transmission of the virus; for instance, airborne dispersal was not considered a major pathway at the beginning of the pandemic, and this inhibited control and containment strategies [8]. Even though thousands of papers have been written on COVID-19 related topics in the past year or so, many questions still remain unanswered, especially in terms of how SARS-CoV-2 transmission is influenced by aspects of the economy, environment, and health. A better understanding of how these factors interact can help us to design early health prevention and control strategies, and to develop better predictive models for public health risk management of SARS-CoV-2 and other novel coronaviruses [9].

This study explores how some of the macro-level drivers of epidemic growth in the United States are associated with COVID-19 case and death doubling times during the first wave of the pandemic (in early 2020). The reason for selecting the United States is not only that it is one of the hardest-hit countries, but also that it provides us with a unique opportunity to study this phenomenon at a macro-scale, since it encompasses a diverse range of climate types over a vast geographical area, with a somewhat homogeneous political system, allowing us to disentangle the effects of the environment from other demographic and socio-economic conditions. Furthermore, the scientific institutions of the United States offer a vast quantity of high-quality data which allows us to investigate our research question rigorously. By focusing on the first wave of the pandemic, it is possible to better isolate the effects of the environment and socio-economic and demographic factors, since it took some time for the population to adopt self-protective behaviors like vaccination, social distancing and mask-wearing; it also took some time for state governments to apply containment measures, like school closures, limits on gathering and non-essential business closures [7, 10, 11].

The empirical strategy for this study relies on county-level aggregated areal health data. The use of aggregate data presents some challenges since we cannot account for individual heterogeneity, which may lead to confounding bias i.e., relationships observed at the group level does not necessarily hold for individuals. Additionally, we may not be able to draw causal inferences due to the likely presence of endogeneity bias (e.g., omitted variable bias, simultaneity, or reverse causality). Nevertheless, this type of empirical investigation maintains high merit, as it enables a quick exploration of geographic associations between the disease and the predictor variables, which can instigate further debate on this topic and may trigger more refined channels of research. The next subsection presents a short analytical framework, explaining how economic, demographic, health, and meteorological factors, as well as containment measures, are expected to influence the spread of the disease, and describes the variables selected to measure such factors.

### 1.1 Analytical framework

SARS-CoV-2 transmission takes place through 4 major pathways including human to human physical contact, indirect contact via fomites, or inhalation of large droplets and fine aerosols [8, 12]. Social distancing can be one of the most effective measures to limit transmission, but this can be rendered ineffective in closed spaces with poor ventilation since the virus can transmit through long-distance airborne dispersal [8, 13, 14]. This study emphasizes socioeconomic, demographic, population health and environmental factors that can influence human to human contact and proximity, and can therefore modulate SARS-CoV-2 transmission [15]. Data on government containment measures will also be analyzed, in part to serve as control variables in the statistical models, but also to provide a description of the early stages of the pandemic in a historical context.

#### 1.1.1 Economic / demographic / health factors

Given the transmission pathways of SARS-CoV-2, as a priori, we would expect to see more infections occur in locations with higher population densities (e.g., metropolitan areas, cities) with high public transport usage, overcrowded living spaces, and industries where business takes place indoors - all of which naturally bring people into closer contact, allowing airborne transmission to take place. To represent this in the models, we select variables representing population density, public transport usage and household overcrowding. We would also expect areas with a higher number of new residents arriving from abroad or out of state, to have had a larger number of outbreaks during the early stages of the pandemic through importation of the virus from infected areas. To represent this in our model, we built a variable that captured the annual rate of new residents arriving to a county from abroad or a different state.

At the beginning of the pandemic, it took some time before a consensus was reached about airborne transmission [2, 8, 12, 13, 16], which had major implications for early policy and practice, like improving ventilation in workspaces and adoption of behavioral changes like mask-wearing. We would expect the adoption of self-protective health behaviors (e.g., social distancing, work from home) that can reduce the chance of contracting and spreading the virus [10, 17, 18] to be harder for low skilled workers or those working in specific economic sectors (like manufacturing). Moreover, the inability to self protect may be accentuated for those who suffer from in-work poverty or precariousness since they may also be obliged to work, even when suffering with symptoms, because of a lack of sick pay, fear of losing a day’s salary and top-down pressures [19–21]. These factors are represented in the empirical models using variables that capture unemployment rates, employment levels in key economic sectors, education of the labor force, and poverty.

Initial reports from the ECDC [22], the WHO [23] and the CDC [24] suggest that those most at risk of serious morbidity and mortality are older people and people with underlying health conditions such as diabetes, obesity, respiratory diseases, cancer, and cardiovascular diseases, poverty is a major risk factor of poor population health and is correlated with such conditions [25–29]. To represent this in the models, variables were selected that capture the age structure of the population, poverty rates, long term air pollution for underlying pulmonary health conditions and the prevalence of diabetes.

#### 1.1.2 Environmental factors

Meteorological factors may affect SARS-CoV-2 transmission by altering human behavior; a basic assumption is people are likely to stay indoors on days with very low or very high temperatures, and/or high rainfall. Furthermore, as a priori, we would expect people to better ventilate their homes/workspaces in places with warmer climates (e.g., leave their windows open, use wall and ceiling fans), which could have an observable overall effect on disease transmission. To represent these factors in the models, variables were selected representing average rainfall, temperate and relative humidity. Meteorological factors can also change the transmission potential and decay rate of the virus in air and on surfaces by altering its stability. [30, 31]. Strong UV light can also inactivate SARS-CoV-2; however, we did not consider this as a strong factor since most transmission takes place indoors [32].

#### 1.1.3 Containment measures

State governments implemented a wide range of measures to tackle COVID-19 outbreaks such as school closures, workplace closures, restrictions on gatherings, close public transport, stay at home requirements, restrictions on internal movement, international travel controls and public information campaigns, all of which could have had some success in suppressing the spread of the disease [33]. To control for this in the models a “Stringency Index” measure was selected that reflects the level of a state government’s response to COVID-19 outbreaks, by quantifying how many measures were implemented and to what degree they were applied.

## 2 Methods

All data were aggregated at the county level, apart from some data on containment measures which are presented at state level. Below a detailed description of the data sources.

### 2.1 Data collection and processing

#### 2.1.1 Morbidity and mortality data

SARS-CoV-2 morbidity and mortality data were sourced from Johns Hopkins University’s Centre for Systems Science and Engineering’s (CSSE) GitHub repository [34]. In general, during the first wave of outbreaks in the US, testing was conducted only on those reporting more serious symptoms (see Additional file 1 - Covid policy tracker). Almost all diagnostic testing for COVID-19 was done with the PCR-based methods, using nasopharyngeal or oropharyngeal specimens (nose or throat swabs).

#### 2.1.2 Economic, demographic and population health data

Data on county population, public transport usage, population age structure, health insurance coverage, immigration, disabilities, and household overcrowding were sourced from the United States Census Bureau using 2015-2019 ACS 5-year estimates [35]. To standardize data across counties, all appropriate variables were converted to percentages/averages of the total county population. A household was considered overcrowded if the number of rooms was less than the number of inhabitants (above 1.01 people per room), this figure included all rooms in a household (not just bedrooms). The disabilities measure captured various health conditions such as difficulty seeing or hearing, restricted movement, learning disabilities, cerebral palsy or other developmental disabilities, or intellectual or mental health disabilities [36].

Population density per km^2^ was calculated using R’s SF package and the United States Census Bureau Cartographic county-level shape-files. Because the range of population density values was very wide, all values above 2500 km^2^ were capped to this value. This modification was tested in the final models and did not affect the results and allowed for better interpretability of the results.

County-level data on unemployment (%), median household income ($), and poverty % were sourced from the USDA Economic Research Service [37]. The “Poverty %” indicator represents the percentage of people/families whose earnings are less than the threshold designated by the Census Bureau’s set of money income thresholds. Data on diabetes prevalence were sourced from the CDC’s diabetes atlas [38]. Economic dependence of a county was represented using the ERS county-level typology data-set [37], this breaks down a county into one of 6 major economic typologies: farming, mining, manufacturing, federal/state government, recreation, and non-specialized.

#### 2.1.3 Environmental data

Temperature (°C), precipitation (1/100”), and relative humidity data were sourced from the Global Surface Summary of the Day (GSOD) data provided by the US National Climatic Data Center (NCDC)[39]. This data-set provides daily GPS observations from all weather stations situated in the US. To join county data with the GSOD weather observations, centroids were created for each county using R’s SF package and the United States Census Bureau’s county shape-files. The K-nearest neighbor join function in R’s SF package was used to create a spatial join between the weather stations (GPS coordinates) and the county centroids. Mean climate values were created for a county-based on data from a maximum of 10 nearest weather stations within a 100km radius of each county centroid.

Data on air quality was sourced from the United States Environmental Protection Agency [40]. We used annual maximum reported Air Quality Index (AQI) values, taken over a 20-year average. This indicator was derived from data from EPA’s AQS (Air Quality System) database. The EPA establishes an AQI based on five major air pollutants including ground-level ozone (*O*_3_), particle pollution (also known as particulate matter, including PM2.5 and PM10), carbon monoxide *CO* sulfur dioxide (*SO*_2_) and nitrogen dioxide (*NO*_2_). The U.S. AQI index runs from 0 to 500. The higher the AQI value, the greater the level of air pollution and the greater the health concern. The AQI is divided into 6 categories, each corresponding to a different level of health concern; generally, they represent 0 to 50 - good; 51 to 100 - moderate; 51 to 100 - unhealthy for sensitive groups; 151 to 200 - unhealthy; 201 to 300 - very unhealthy; 301 and higher - hazardous.

#### 2.1.4 Containment measures

Data on county-level stay-at-home orders (lock-down) were extracted from the CDC’s “U.S. State, Territorial, and County Stay-At-Home Orders’ ’ dataset [41]. This dataset provides information on county-level executive orders, administrative orders, resolutions, and proclamations and can be used to determine the date of county-level stay-at-home orders (lock-down).

Data on state-level control measures were sourced from the Oxford COVID-19 Government Response Tracker (OxCGRT) data set [42]. We used the “Stringency Index” variable from this dataset to account for the application of state-level control measures in our final models. The composite time-series measure, ranging from 0 to 100 (100 = strictest) is based on 9 response indicators including data on school closing, workplace closing, restrictions on gatherings, close public transport, stay at home requirements, restrictions on internal movement, international travel controls, and public information campaigns. The indicator reflects the level of a state government’s response to COVID-19 outbreaks and quantifies how many measures were implemented, and to what degree they were implemented. The index cannot ascertain whether a government’s policy has been implemented effectively nor the effectiveness of an individual measure [33]. To get an estimate of a government’s response leading up to the first lock-down (compulsory stay at home order), the average stringency index value was calculated using a time window: from the day the first 5 cases were reported the day before the first lock-down. Arkansas, Iowa, Nebraska, North Dakota, and South Dakota did not implement state-wide lockdowns. In these states, we took the average score from the day the first cases were reported to the last lock-down date in our sample (2020-07-04) to make this value comparable to other states.

### 2.2 Study design

The spread of the disease (epidemic growth) is modeled by calculating COVID-19 case and death doubling times; these measures are then used as dependent variables to explore associations between epidemic growth, socio-economic, demographic, and environmental factors, and population health. Doubling times capture exponential growth, in this instance, the number of days taken for cases and deaths to double. This measure has several advantages: first, it provides a way for standardizing differences in sampling effort between different locations and health authorities; second, because it provides us with a time determinant measure to facilitate understanding of the spread of the virus. In other words, this metric not only has the advantage of accounting for population size but also incorporates a time dimension. Therefore, COVID-19 transmission is measured by calculating doubling times for infections and mortality, at the county level [43–46].

#### 2.2.1 Calculating case and death doubling times

Doubling times are calculated by capturing a window of infection opportunity i.e., from the time the first cases/deaths were detected in a county, to the first major state or county level intervention, such as a compulsory stay-at-home order, otherwise known as a lock-down (see Figure 1). We also accounted for disease progression, since there is a lag between the time an event is reported (a case or death) and the date the transmission event took place. We lag all case and mortality data by the maximum incubation period (onset of symptoms) or maximum time from final infection to death.

**Figure 1.**
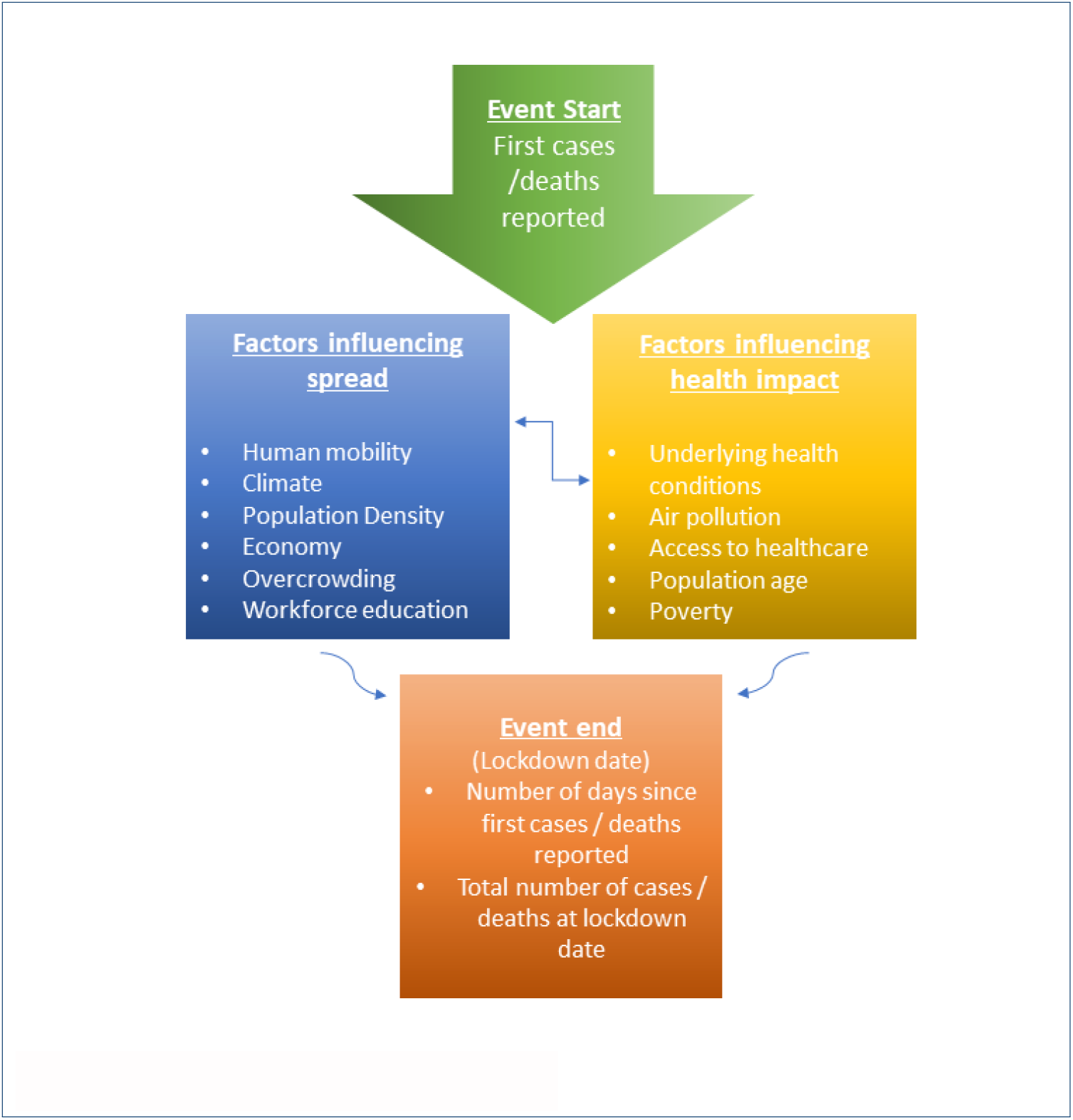
Study design: capturing epidemic growth

#### 2.2.2 Case data

For data on confirmed COVID-19 cases, we set a lag of 21 days, which considers a maximum 14-day incubation period based on estimates by Lauer 2020 [47], with an extra 7 days to account for any reporting delays. We, therefore, use case data to calculate the doubling times for any dates up to 21 days post-lock-down. The count was started when the county reached 50 confirmed cases, over a minimum of 7 days.

#### 2.2.3 Mortality data

For the mortality data set, we set a lag of 42 days which includes the maximum 14-day incubation period based on estimates reported by Lauer et al., 2020 [47] and a maximum of 21 days from the first onset of symptoms to death as reported by Verity et al., 2020 [48], plus an extra 7 days to account for any reporting delays. Since the mortality data-set contained fewer observations than the cases data set, we started the count when the county reported 20 deaths over at least 7 days. This value yields enough observations to carry out the study, although the doubling times may be less stable than that of the case data-set.

To calculate the case and death doubling times for each county, we used the following formulas:

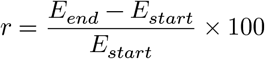

Where:

*r* = growth rate;

*E*_*start*_ = Start of the event - when the 50 cases / 20 deaths are detected

*E*_*end*_ = End of the event - cumulative cases / deaths per county at the lock-down date;

Next, the doubling time is calculated using the following formula:

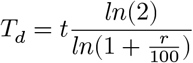

Where:

*T*_*d*_ = doubling time in days

*t* = time in days (Estart to Eend)

*r* = growth rate

Arkansas, Iowa, Nebraska, North Dakota, and South Dakota did not implement a state-wide lock-down (stay at home order), so an artificial date was set to calculate doubling times, mirroring the latest lock-down date in our sample (2020-07-04).

#### 2.2.4 General additive regression model to assess the impact of independent variables on doubling times at the county level

One of the main issues with our data-set is that it did not meet some basic assumptions for statistical inference, that is the data are not independent and identically distributed random variables (iid). More specifically, observations cannot be considered independent because of spillover effects from neighboring regions, therefore we needed to implement an appropriate statistical design to control for spatial pseudoreplication (lack of independence). We could deal with this in two ways, 1) either relaxing the assumption of independence and estimating the spatial correlation between residuals, or 2) model the spatial dependence in the systematic part of the model [49]. We opted to use a Generalized Additive Model (GAM) using R’s Mgcv statistical package because of its versatility and ability to fit complex models that would converge even with low numbers of observations and could capture potential complex non-linear relationships. One of the advantages of GAMs is that we do not need to determine the functional form of the relationship beforehand. In general, such models transform the mean response to an additive form so that additive components are smooth functions (e.g., splines) of the covariates, in which functions themselves are expressed as basis-function expansions. The spatial auto-correlation in the GAM was approximated by a Markov random field (MRF) smoother, which represents the spatial dependence structure in the data. We used R’s Spdep package to create a queen neighbors list (adjacency matrix) based on counties with contiguous boundaries i.e., those sharing one or more boundary points. The local Markov property assumes that a region is conditionally independent of all other regions unless regions share a boundary. This feature allowed us to model the correlation between geographical neighbors and smooth over contiguous spatial areas, summarizing the trend of the response variable as a function of the predictors [50]. Models were fit using a gamma distribution, after inspecting our data, we concluded that the gamma distribution worked well with the shape of our response variable, which was positively skewed (i.e., non-normal, with a long tail on the right). The gamma distribution is a two-parameter distribution, where the parameters are traditionally known as shape and rate. Its density function is:

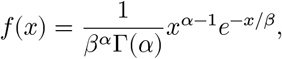

where *α* is the shape parameter and *β*–1 is the rate parameter (alternatively, *β* is known as the scale parameter).

The empirical model can then be written as:

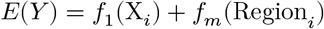

Where the *f*(·) stands for smooth functions; *E*(*Y*)_*i*_ is equal to infection or death doubling time in region *i*, which we assume to be gamma-distributed; *Xi* - is a vector of economic, demographic, environmental and climate variables (as described in the previous section). *Region*_*i*_ represents neighborhood structure of the region.

Analysis of model diagnostic tests didn’t reveal any major issues, in general residuals appeared to be randomly distributed. For robustness, models were also fit using the Gaussian and Tweedie distributions, and also fit using a non-additive-GLM (see Additional file 2).

## 3 Results

To carry out the empirical analysis, a unique spatial data-set was compiled that captured potential drivers of human-to-human SARS-CoV-2 transmission and risk factors of serious infections and mortality due to COVID-19 in US counties.

### 3.0.1 Descriptive statistics

Two sources of information were analyzed, data on confirmed cases and deaths. Tables 1 and 2 provide summary statistics for our final data-sets.

**Table 1.** Cases data-set - summary statistics

| Statistic | N | Min | Max | Mean | St. Dev. |
| --- | --- | --- | --- | --- | --- |
| Confirmed cases | 640 | 54 | 35,571 | 950.9 | 2,953.7 |
| Incidence per 100,000 | 640 | 15.8 | 5,611.3 | 277.5 | 434.1 |
| Study period days | 640 | 7 | 40 | 18.7 | 7.5 |
| Growth rate (over period) | 640 | 8 | 62,305 | 1,503.5 | 5,040.1 |
| Cases doubling time | 640 | 2.5 | 72.0 | 11.6 | 8.4 |
| County population | 640 | 5,861 | 10,081,570 | 373,339.6 | 657,728.9 |
| Pop density per km2 | 640 | 2.0 | 2,500.0 | 245.3 | 415.8 |
| Stringency index | 640 | 11.0 | 64.1 | 34.1 | 10.5 |
| Public transport usage (pop %) | 640 | 0.0 | 32.0 | 1.3 | 3.1 |
| Median household income (\$) | 640 | 26,348 | 151,806 | 65,642.1 | 18,915.3 |
| Unemployment % | 640 | 1.8 | 12.0 | 3.8 | 1.2 |
| Pop % 65+ | 640 | 7.9 | 41.1 | 16.5 | 4.2 |
| Poverty % | 640 | 2.7 | 36.6 | 13.1 | 5.9 |
| Health insurance coverage % | 640 | 62.2 | 99.9 | 98.0 | 3.0 |
| Annual new arrivals / by pop (%) | 640 | 0.0 | 3.1 | 0.5 | 0.4 |
| Pop % with disabilities | 640 | 5.0 | 25.7 | 13.1 | 3.3 |
| Pop % with diabetes | 640 | 2.2 | 23.1 | 10.1 | 2.9 |
| Household overcrowding % | 640 | 0.1 | 7.0 | 1.3 | 1.0 |
| Pop % with degree or higher | 640 | 3.6 | 29.0 | 12.6 | 4.6 |
| Temperature (°C) | 640 | −3.4 | 24.7 | 12.7 | 6.4 |
| Precipitation (1/100") | 640 | 0.0 | 9.2 | 2.1 | 1.6 |
| Relative humidity | 640 | 30.7 | 86.1 | 67.0 | 8.6 |
| Max AQI | 640 | 32.5 | 347.4 | 130.4 | 31.3 |

**Table 2.** Mortaility dataset - summary statistics

| Statistic | N | Min | Max | Mean | St. Dev. |
| --- | --- | --- | --- | --- | --- |
| Confirmed cases | 263 | 22 | 4,902 | 233.4 | 541.3 |
| Incidence per 100,000 | 263 | 1.4 | 284.6 | 39.9 | 45.4 |
| Study period days | 263 | 7 | 56 | 24.4 | 10.2 |
| Growth rate (over period) | 263 | 9.5 | 24,410.0 | 906.5 | 2,346.6 |
| Cases doubling time | 263 | 5.7 | 80.0 | 18.5 | 12.5 |
| County population | 263 | 8,737 | 10,081,570 | 677,179.5 | 929,606.3 |
| Pop density per km2 | 263 | 2.9 | 2,500.0 | 442.2 | 570.3 |
| Stringency index | 263 | 11.0 | 64.1 | 32.4 | 11.6 |
| Public transport usage (pop %) | 263 | 0.0 | 32.0 | 2.4 | 4.5 |
| Median household income (\$) | 263 | 36,894 | 151,806 | 71,804.3 | 20,582.3 |
| Unemployment % | 263 | 1.8 | 9.6 | 3.7 | 1.0 |
| Pop % 65+ | 263 | 9.5 | 41.1 | 16.1 | 4.0 |
| Poverty % | 263 | 2.7 | 30.7 | 12.0 | 5.1 |
| Health insurance coverage % | 263 | 89.9 | 99.6 | 98.7 | 1.2 |
| Annual new arrivals / by pop (%) | 263 | 0.01 | 2.2 | 0.6 | 0.4 |
| Pop % with disabilities | 263 | 5.8 | 23.5 | 12.1 | 2.9 |
| Pop % with diabetes | 263 | 5.2 | 22.3 | 9.3 | 2.3 |
| Household overcrowding % | 263 | 0 | 6 | 1.4 | 1.0 |
| Pop % with degree or higher | 263 | 3.9 | 27.6 | 14.2 | 4.4 |
| Temperature (°C) | 263 | 0.2 | 25.3 | 11.6 | 5.9 |
| Precipitation (1/100") | 263 | 0.0 | 7.4 | 2.0 | 1.4 |
| Relative humidity | 263 | 25.5 | 81.6 | 64.9 | 8.8 |
| Max AQI | 263 | 41.5 | 347.4 | 143.8 | 31.5 |

To calculate doubling times, counties were only selected that had reported at least 50 cases or 20 deaths over a minimum 7-day period before the first lock-down. Both sources of information were chosen as they allow us to explore and compare different features and characteristics of the epidemic. Figures 1 and 2 map the geographical distribution for case and death doubling times in counties that met our inclusion criteria (colored from red to yellow). Major cities with populations > 250,000 people are highlighted on each map. The counties first affected by SARS-CoV-2 during the first wave of the epidemic tended to be located around major cities and metropolitan areas on the east coast, mid west, and south of the United States, with high population density and presumably higher numbers of international and domestic travelers.

**Figure 2.**
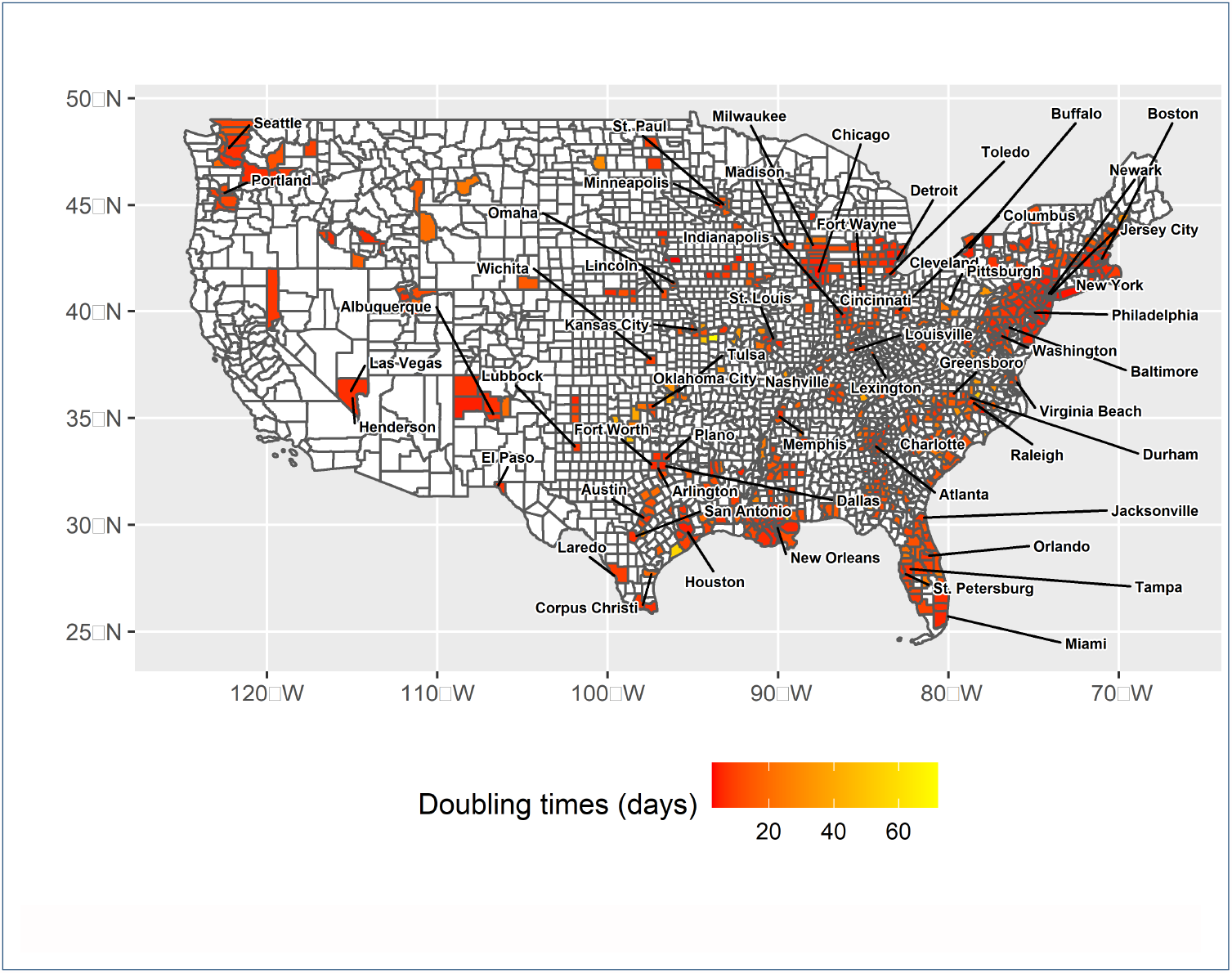
Covid-19 case doubling times in US Counties (Data source: Johns Hopkins University)

### 3.0.2 Regression results

It was not possible to explore the individual impact of all the variables in our data-set because of collinearity issues (see Additional file 2). Public transport was positively correlated with population density so therefore removed from the analysis. Median income was also removed from the analysis because it was positively correlated with education, and negatively correlated with poverty, disabilities and diabetes.

Tables 3 and 4 show the results of the statistical analysis for both data sets and summarize the relevant statistics (AIC, Deviance, Adjusted R squared (*R*^2^ and so on) to compare the different specifications. Both statistical models were built in a step-wise fashion using the lowest Akaike Information Criterion (AIC) and *R*^2^ to help us assess the different specifications. Variables were included in each specification according to their category i.e., spatial, socio-economic, and environmental. All variables were included in the final specification to ascertain the contribution of each driver or risk factor, all else equal. Note that, as we are not estimating a standard regression model, the figures reported should not be read as coefficients, but degrees of freedom of the smooth terms. Given that we cannot interpret the coefficients to infer the sign and magnitude of the relationship, we visualize it by plot. Figures 3-11 plot the partial effects—the relationship between a change in each of the covariates and a change in the fitted values in the full model. Standard errors on the plots show the 95% confidence interval for the mean shape of the effect.

### 3.0.3 Case data model

Table 3 and figures 3-7 show the results of the model fit using case data. The “Spatial” model was fit first to estimate the contribution of the spatial lag component against the other specifications. A high proportion of the variance is explained just by controlling for spatial correlation between counties (*R*^2^ 0.35). The “Full model” has the best fit in terms of the AIC and adjusted *R*^2^, followed by the socio-economic model, and finally the environmental model. The adjusted *R*^2^ in the final model is 0.56, indicating that 56% of the variance in our model is explained by the explanatory variables.

**Table 3.**
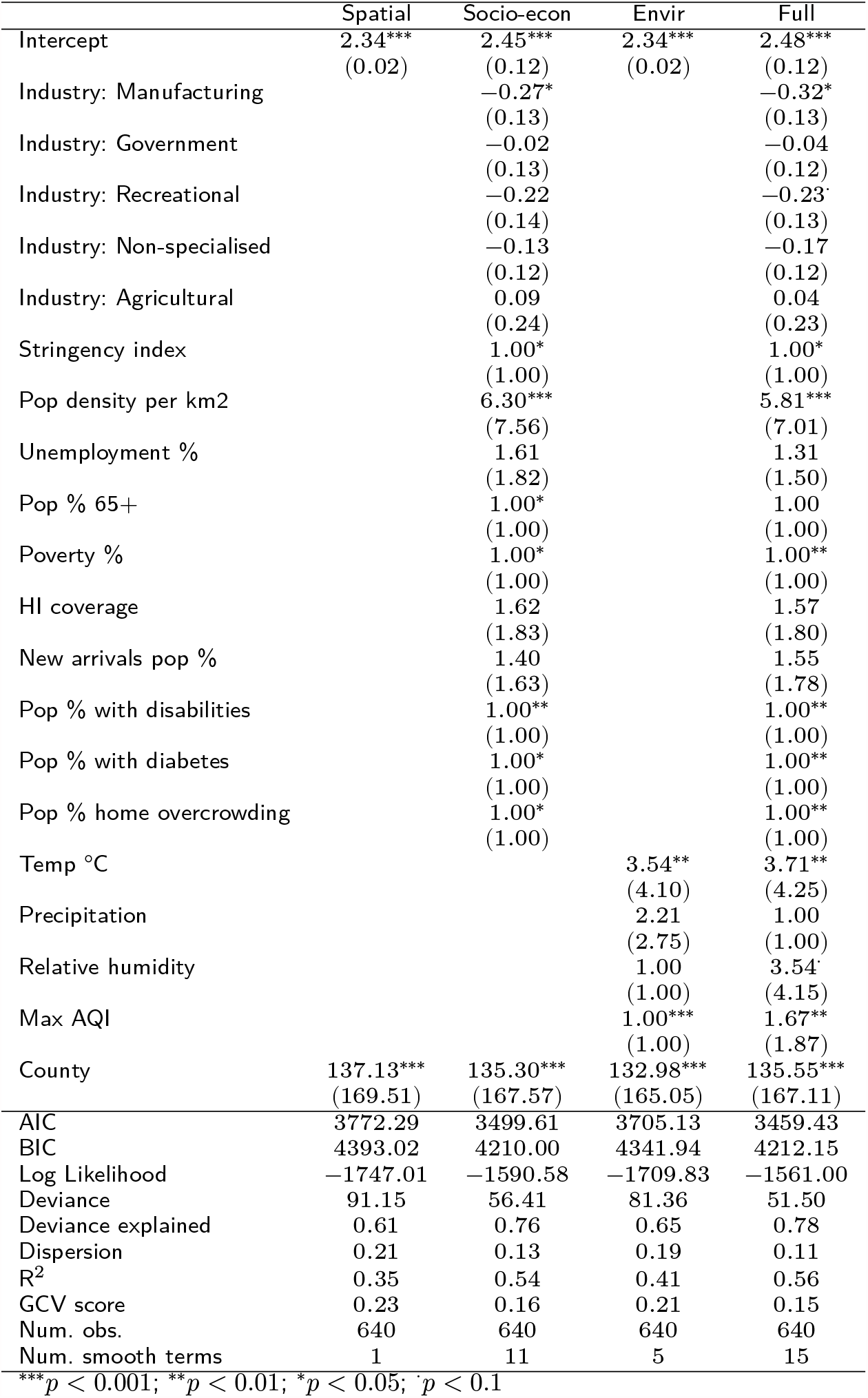
COVID-19 Infection model

**Figure 3.**
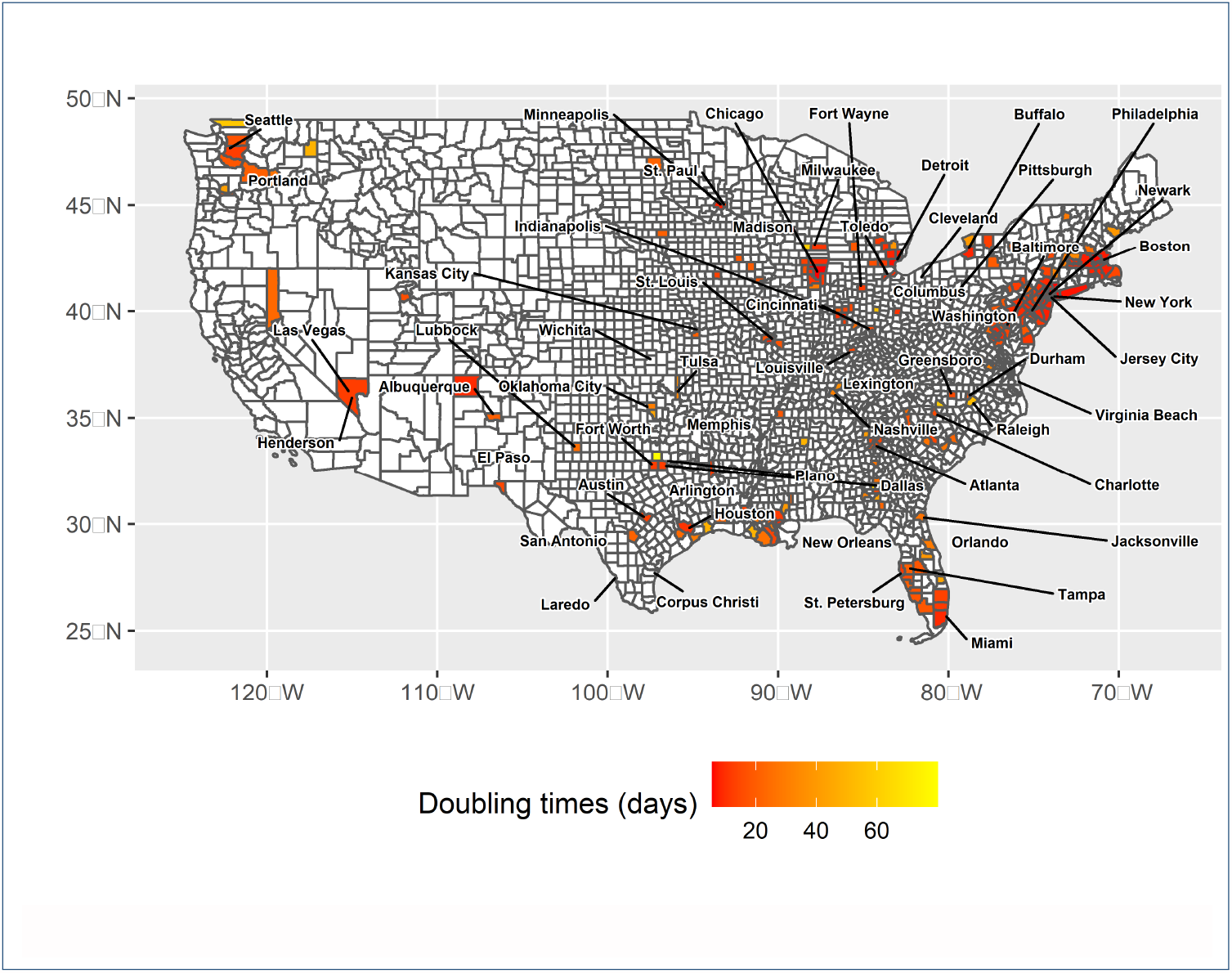
Covid-19 death doubling times in US Counties (Data source: Johns Hopkins University)

**Figure 4.**
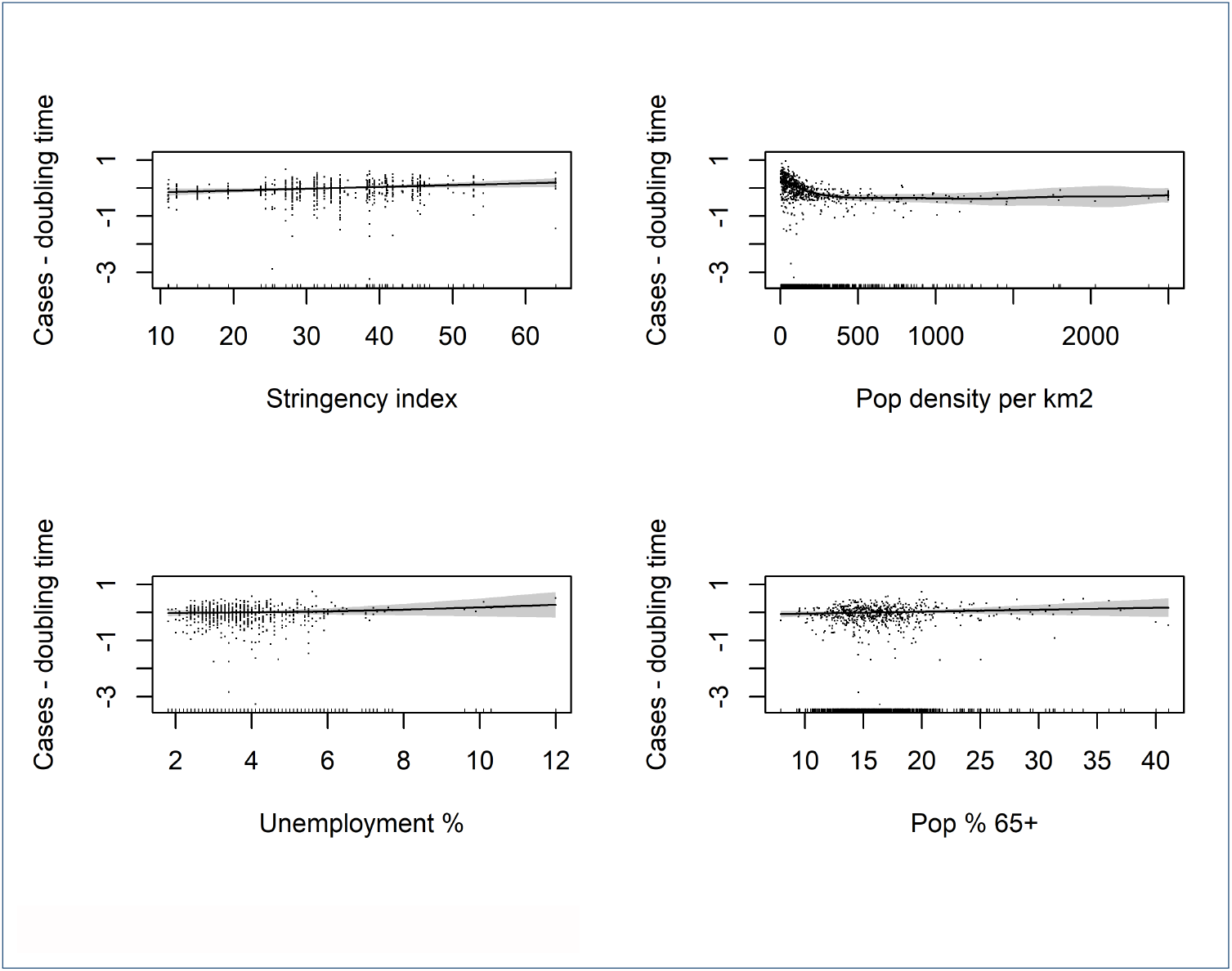
Partial effects plots 1: Case model

**Figure 5.**
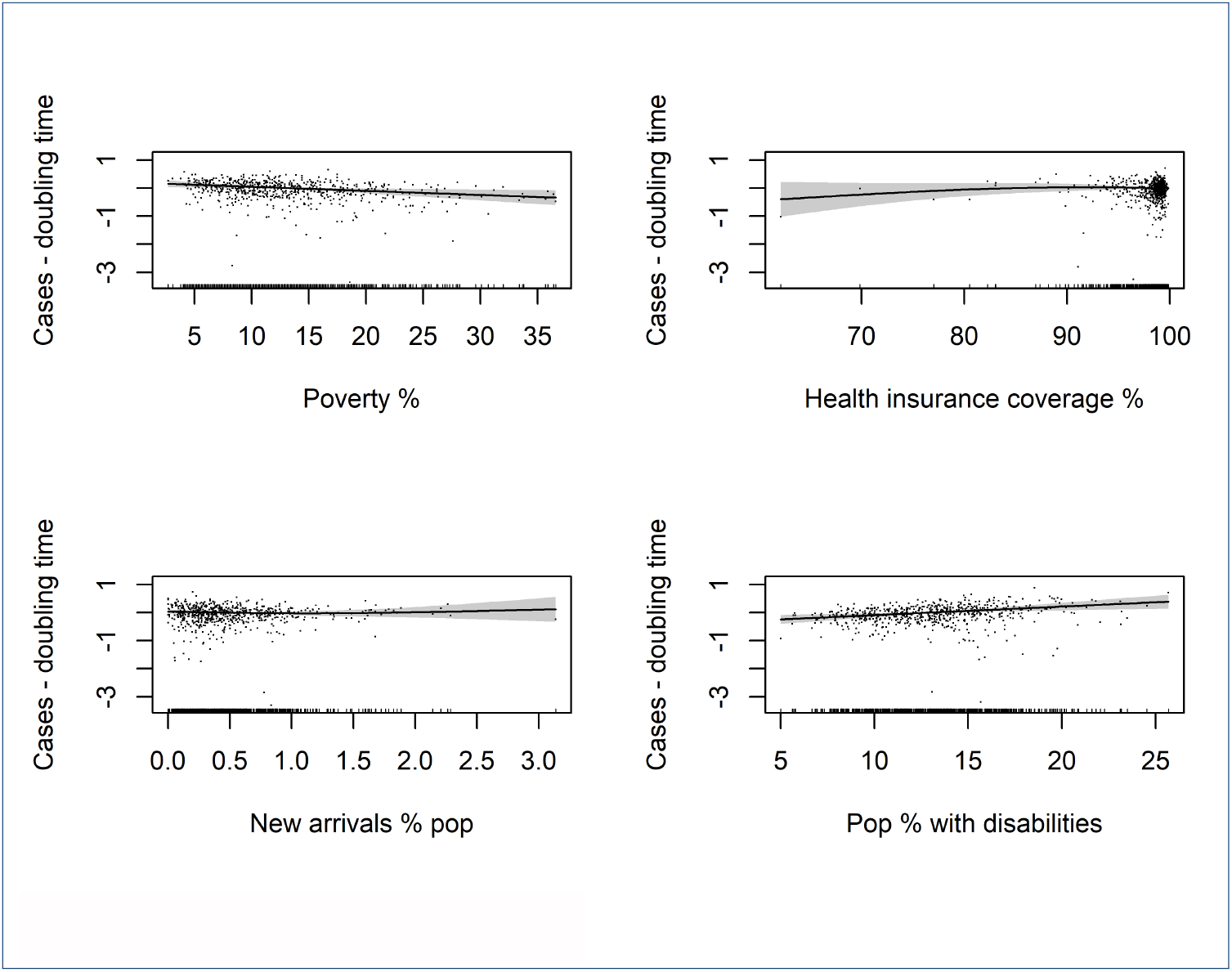
Partial effects plots 2: Case model

**Figure 6.**
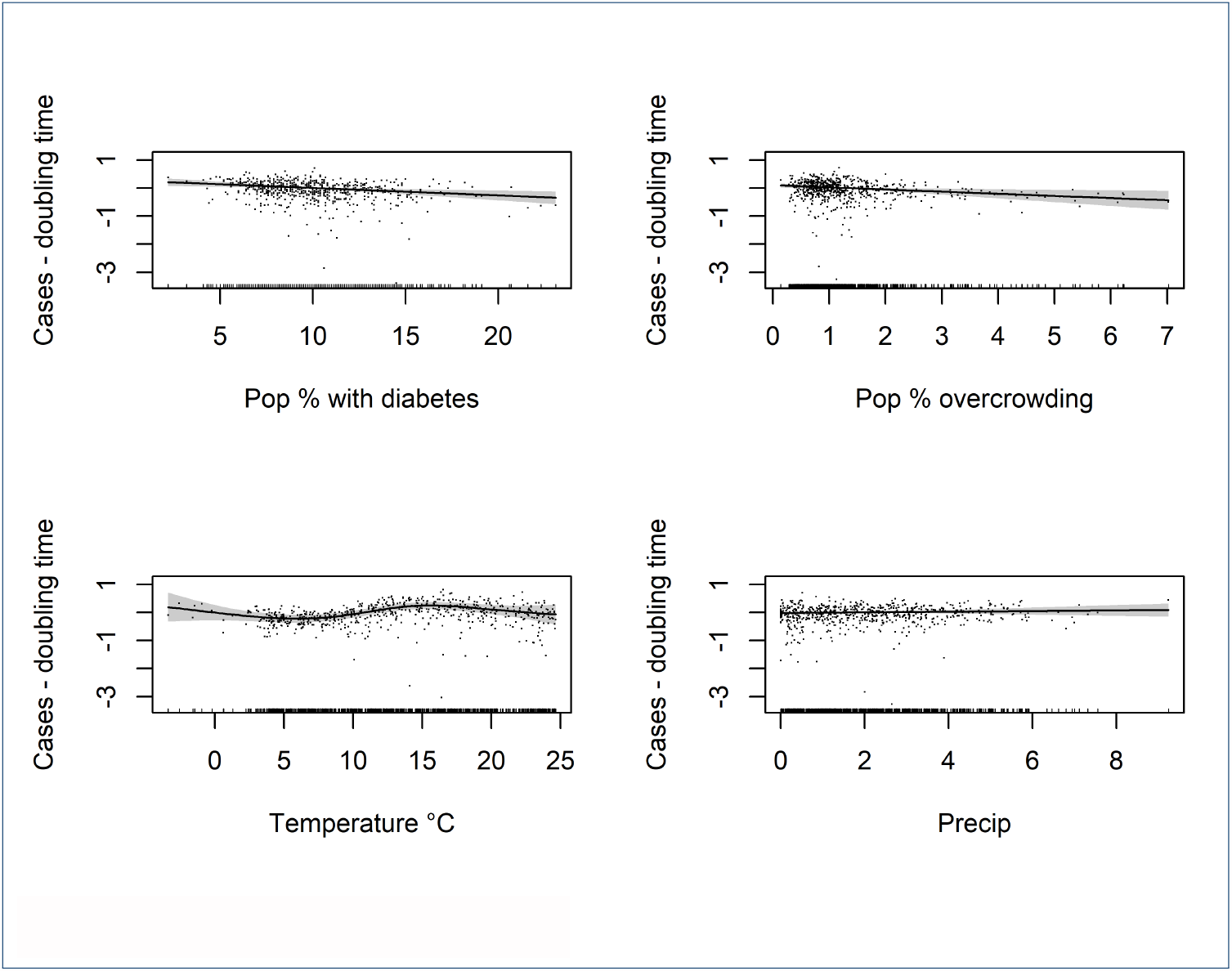
Partial effects plots 3: Case model

**Figure 7.**
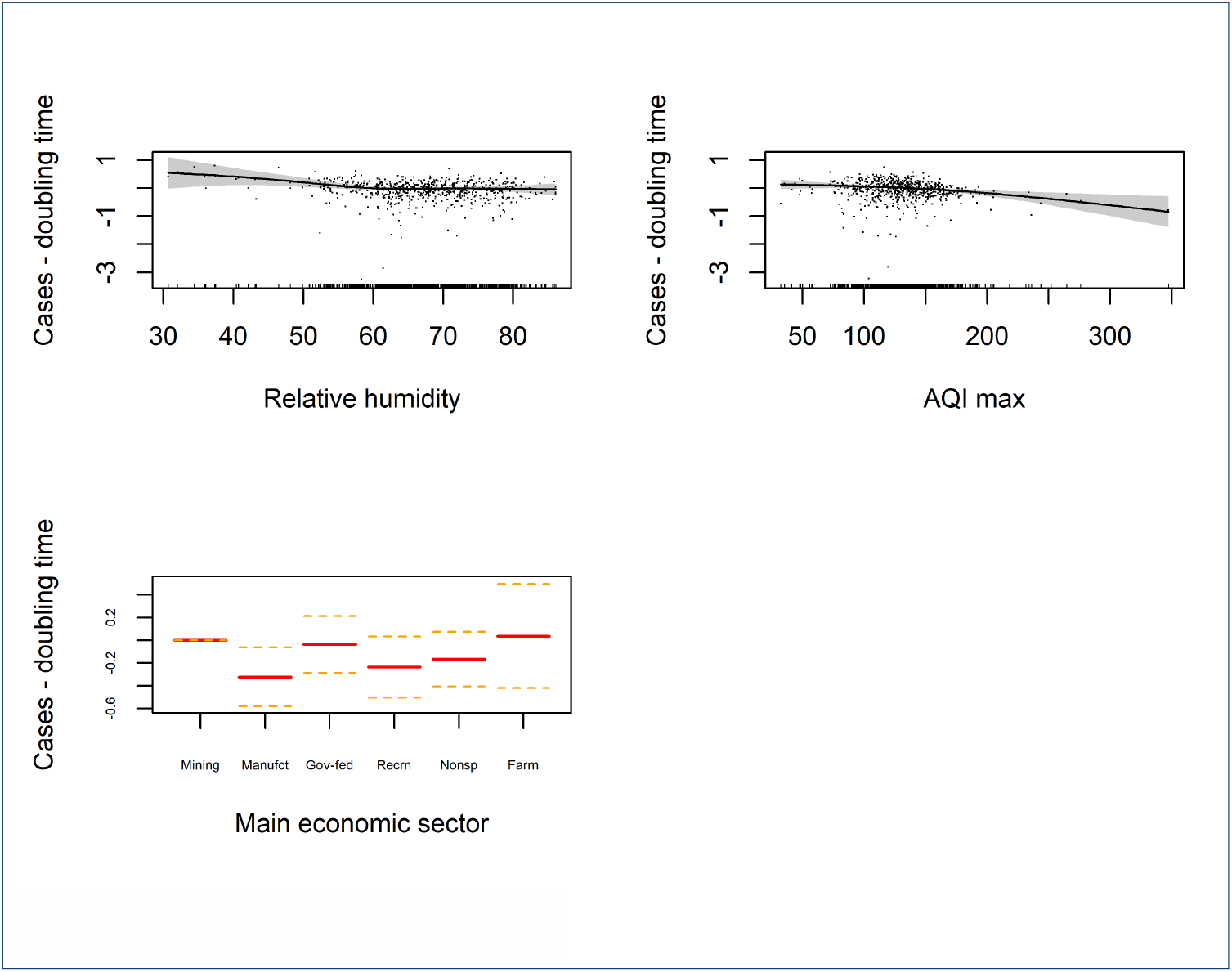
Partial effects plots 4: Case model

As for the contribution of individual variables on case doubling times, counties with manufacturing and recreation as their predominant economic activity were associated with faster case doubling times. The stringency index variable, which captures the number of containment measures adopted by states, and the degree to which they were implemented, is also statistically significant (p < 0.05), and has a positive relationship with the case doubling times, suggesting that measures had some success in suppressing the virus. Human population density per *km*^2^ is highly significant (p < 0.001), higher densities are associated with faster case doubling times, although the relationship is not linear and flattens out at higher population densities. Although the slope is gentle, “Poverty %” is a highly significant (p < 0.01) predictor of case doubling times, the relationship is negative which means doubling times decrease with higher levels of poverty (in other words, the infection spreads faster). On the contrary, the variable “Pop % with disabilities” (p < 0.01) is associated with slower case doubling times. The prevalence of diabetes (Pop % with diabetes) in a county, an indicator that not only represents the disease itself, but also a range of other conditions such as obesity, poor diet, lack of exercise was also a significant (p < 0.01) predictor of faster case doubling times. “Population % home overcrowding”, which represents the percentage of households in a county where there is less than one room per inhabitant (> 1.01 people per room) is highly significant (< 0.01) and is associated with faster case doubling times. Temperature is also a good predictor of case doubling times; higher temperatures appear to slow case doubling times. (p < 0.01). Although this relationship was not entirely clear, some counties with low temperatures also seemed to have slower doubling times. Although the confidence intervals are much wider and at lower temperatures, meaning the results are less accurate. “Max AQI”, which represents the maximum air quality index values averaged over 20 years, is also highly significant; case doubling times are faster in locations with poor air quality (p < 0.01).

### 3.0.4 Mortality model

Table 4 and figures 8-11 show the results of our model fit using mortality data. A high proportion of the variance is explained just by controlling for spatial correlation between counties (*R*^2^ 0.22). The “Full model” has the best fit in terms of the AIC and adjusted *R*^2^ 0.48, followed by the socio-economic model (0.44) and the environmental model (0.31). The “Stringency index” indicator is statistically significant (p < 0.05) and has a positive relationship with death doubling times; more stringent containment measures appeared to suppress COVID-19 mortality. “Population density per *km*^2^” (< 0.001) is also an important predictor: generally, higher population density is associated with faster death doubling times, however, this trend reverses at around 1400 inhabitants per km^2^ and levels off. “Population % 65+” (< 0.001) is highly significant; higher values are associated with faster death doubling times. Again, as with the case data analysis, “Poverty %” is also a highly significant predictor of death doubling times (< 0.001). “Pop % with disabilities” (< 0.01) is also highly significant; like with the case data model, this predictor is positively associated with slower death doubling times. The prevalence of diabetes (Pop % with diabetes) in a county is also a significant predictor (p < 0.05) of faster death doubling times, as is the air quality index (“Max AQI”), which is highly statistically significant (p < 0.01). Temperature and precipitation are slightly significant (p < 0.1). and appear to slow down death doubling times at higher values.

**Table 4.**
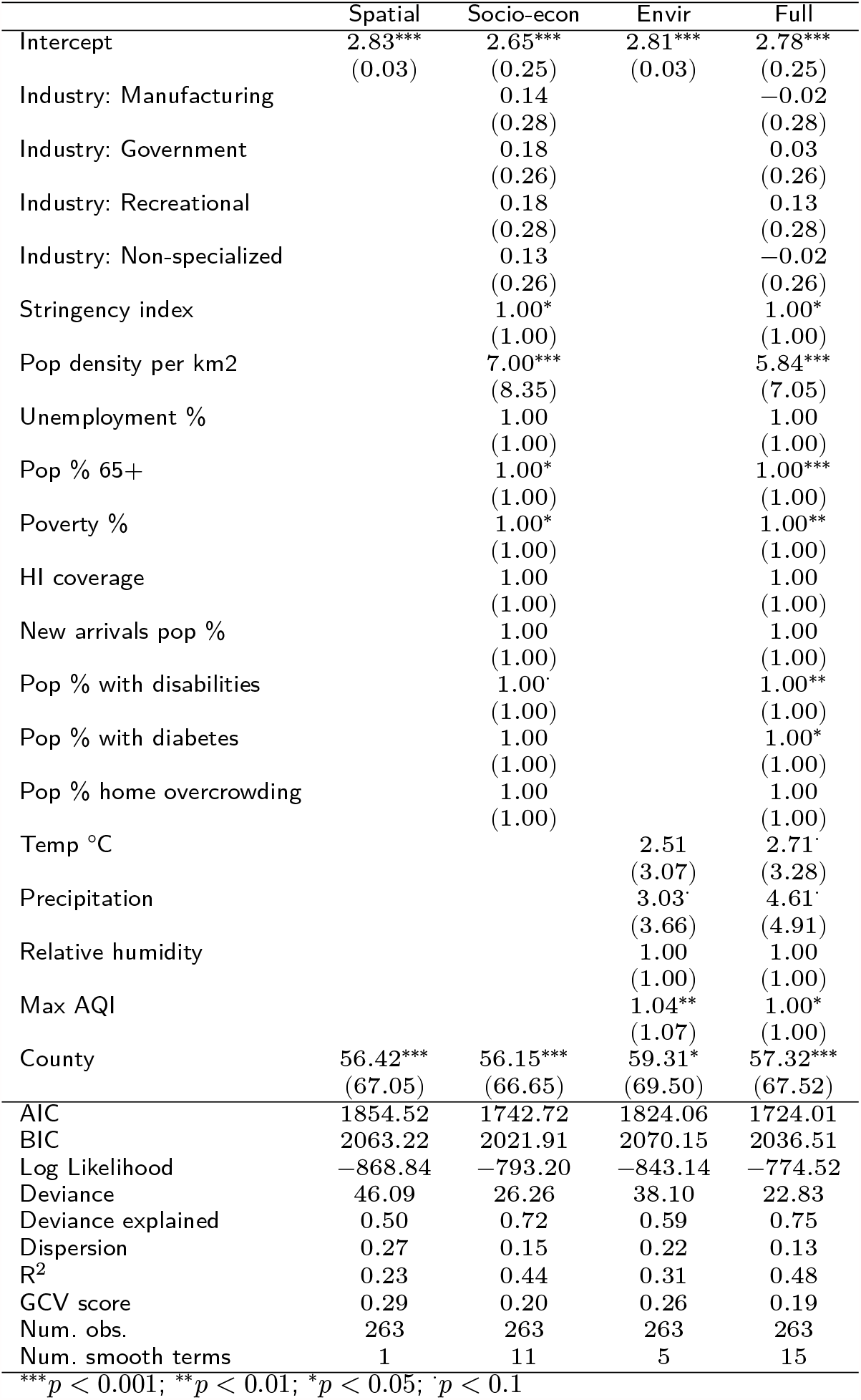
COVID-19 mortality model

**Figure 8.**
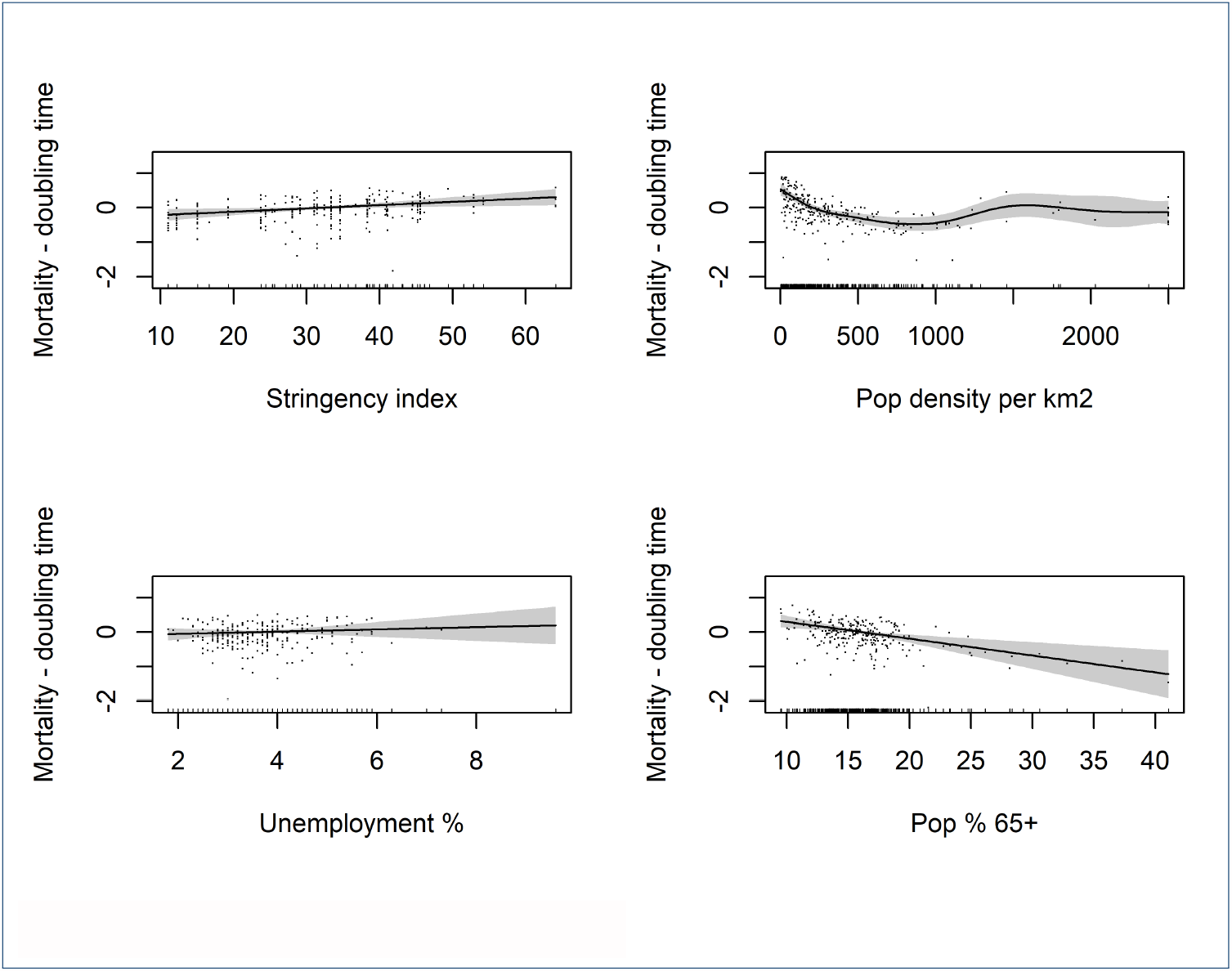
Partial effects plots 1: Mortality model

**Figure 9.**
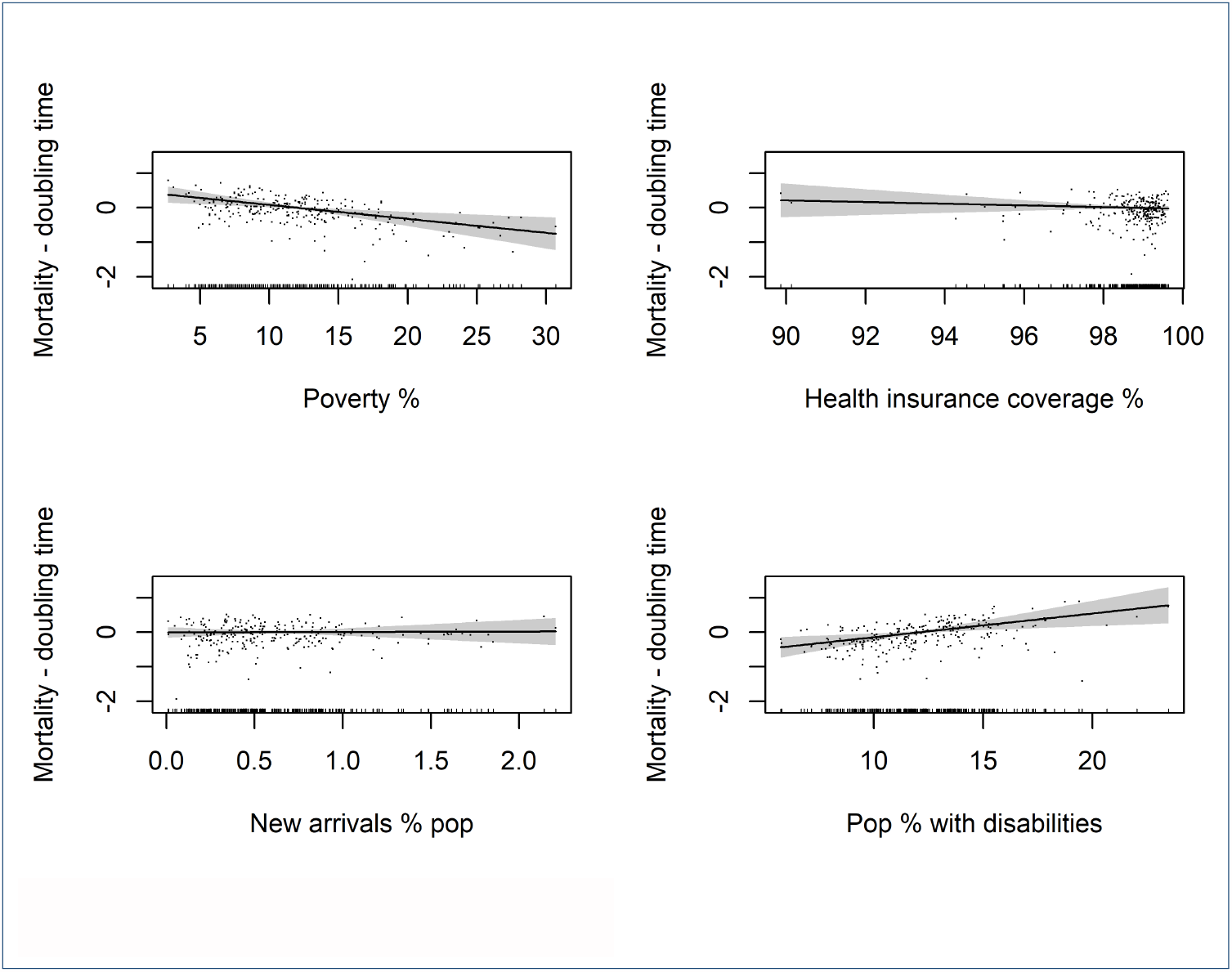
Partial effects plots 2: Mortality model

**Figure 10.**
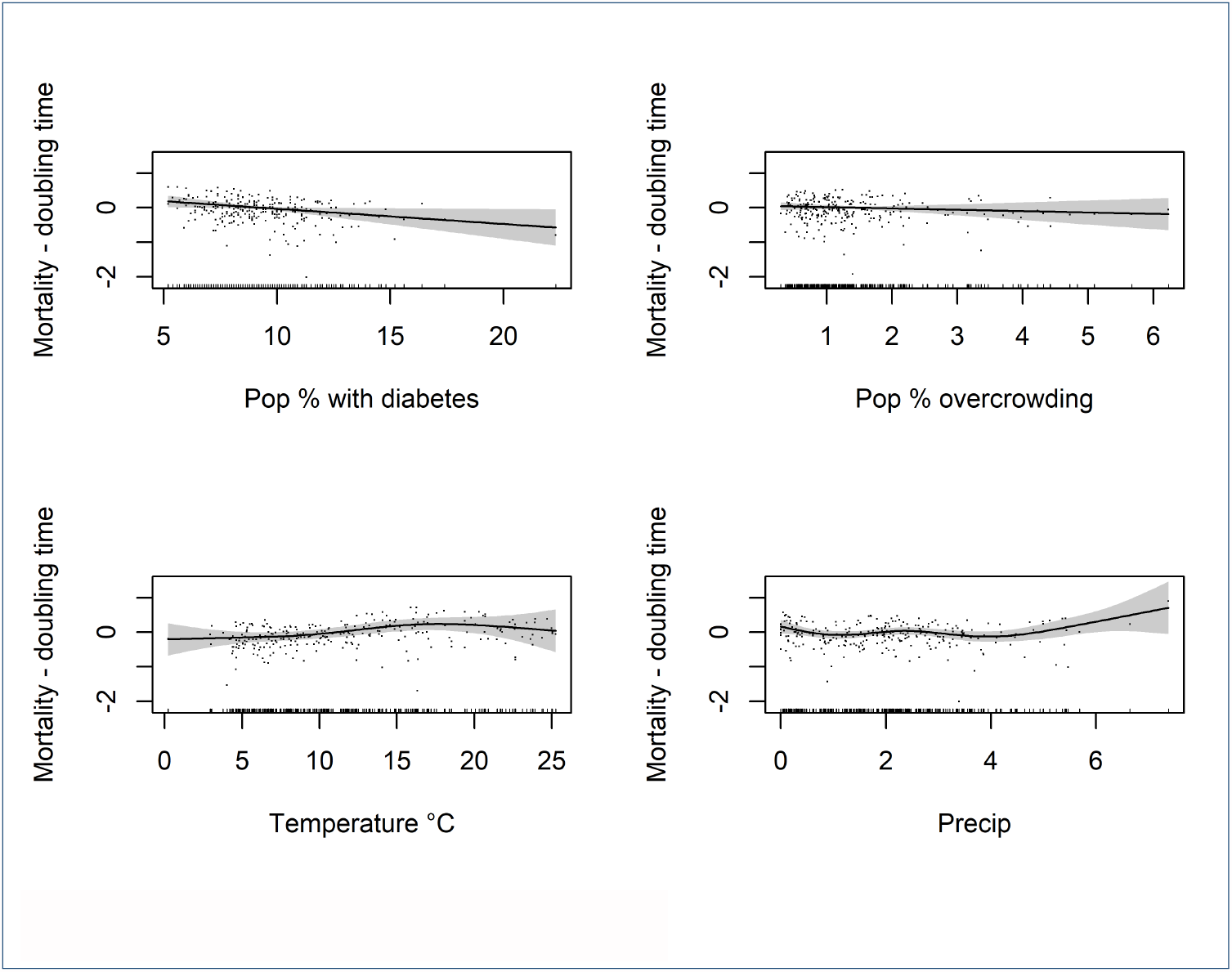
Partial effects plots 3: Mortality model

**Figure 11.**
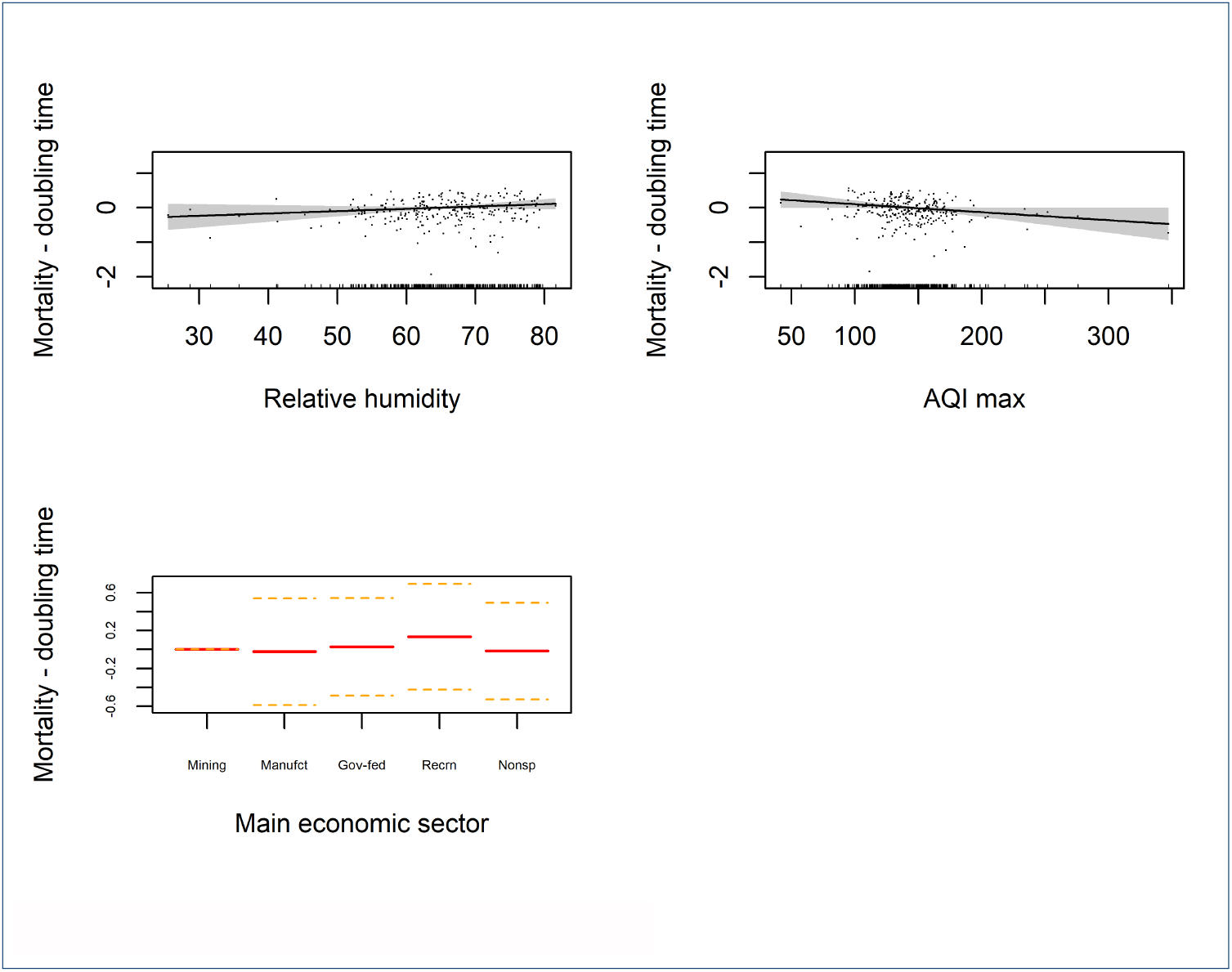
Partial effects plots 4: Mortality model

## 4 Discussion

In this study, we examined which socio-economic, demographic, and environmental factors are associated with SARS-CoV-2 epidemic growth. To explain biases in reporting, we included health risk factors that can contribute to serious SARS CoV-2 infections and deaths. We would expect case reporting to be a function of all these factors since testing policy during this phase of the epidemic was aimed at those with symptoms (see Additional file 1 - COVID-19 policy tracker).

We can also assume that, during this wave of the epidemic in the US, only one strain of SARS-CoV-2 (although always evolving) was in circulation and therefore the variation in infection and death rates across space can be attributed to external factors i.e., testing differences, aspects of the population and environment, rather than variation in viral traits/strains. Furthermore, no vaccines were yet in circulation.

### 4.1 Containment measures to reduce disease spread

During the first wave of the epidemic in the US, governments, and public health systems were initially caught off guard by the rapid spread of the virus. Some of the states did apply more rigorous control measures than others, attempting to suppress the spread of the virus early on e.g., by restricting gatherings, closure of public spaces, creating public awareness campaigns and contact tracing (see Additional file 1). Stringency index scores in both our models are associated with slower doubling times and can be interpreted as, the more stringent the measures applied by state governments early on, the more success they had in suppressing the virus.

### 4.2 Socio-economic, economic, and demographic factors

Results show that human population density is one of the strongest predictors of case and death doubling times, the relationship is negatively linear to a point, where higher population densities are associated with faster doubling times, but this trend tends to level off at population densities of above 400 people per *km*^2^, and reverses slightly for death doubling times at densities above 1000 people per *km*^2^. Perhaps because of features relating to the built environment i.e. building types, age structures, demographic or socio-economic conditions associated with wealthier city dwellers. However, in general, the relationship between population density and COVID-19 transmission is logical given the virus mainly transmits when humans are in close proximity to one another. Human population density also captures other important features of the built environment; for example, locations with high population density are cities or metropolitan areas, usually with high public transport usage, more recreational businesses like restaurants and bars, and indoor work-spaces like offices. All of which naturally bring people into closer contact and encourages airborne transmission of SARS-CoV-2.

Results also show that counties that rely on manufacturing or recreation as their main economic activity, also tend to have faster case doubling times. Again, this is likely due to aspects of the work environment like the lack of proper physical distancing and ventilation. These findings are corroborated by studies [51, 52] that report many SARS-CoV-2 clusters were linked to a variety of indoor settings including households, hospitals, elderly care homes, and food processing plants (classed as factories). This concept is also further supported by our indicator representing household overcrowding, which is another strong predictor of case reporting doubling time. However, these variables are only significant in the infections model and not the mortality model. One possible explanation is that they represent transmission among younger people of working age, students, and younger families, who are less likely to die from COVID-19.

In terms of age population structure, having a higher proportion over 65-year-old’s was also a significant predictor of faster death doubling times, concurrent with the literature and common understanding about the disease; age is one of the major risk factors. Major outbreaks have occurred in care homes [51] suggesting that some of the counties most affected by COVID-19 in the first wave of the epidemic was in locations with a higher proportion of retirees and care homes.

In terms of other socio-economic factors affecting the disease, poverty was also a significant predictor of faster doubling times in both case and mortality models. As mentioned in the conceptual framework, this can be explained since those who suffer from in-work poverty are likely to be doing jobs where it is difficult to work from home or adopt self-protective health behaviors such as social distancing [10]. Furthermore, even when suffering from symptoms, many low skilled workers and precarious workers may have been obliged to work because of a lack of sick pay, fear of losing a day’s salary and pressures from bosses [19–21]. Poverty is also a risk factor of poor population health and is correlated with a multitude of underlying health conditions believed to lead to adverse outcomes for those suffering from COVID-19 [25]. This is further supported by the results of our final models; higher diabetes prevalence is also associated with faster case and death doubling times. Again, those suffering from diabetes are likely to suffer from comorbidities such as obesity and heart problems [53]. These results are also concurrent with work conducted by Williamson et al., 2020 [54], who found that greater age, deprivation, diabetes, severe asthma, and various other medical conditions were at higher risk of death due to COVID-19 infection. For both data-sets “Pop % with disabilities” tended to be correlated with slower doubling times. Although this group may be vulnerable to COVID-19 infections, they can often suffer from social isolation which provides some explanation. Furthermore, these groups are more likely to self-isolate [55, 56] to avoid infections.

#### 4.2.1 Environmental factors

Although a broad measure, the air quality index (“Max AQI”) provides us with a way to proxy for counties with poor air quality and population-level pulmonary health conditions, caused by long-term exposure to harmful pollutants such as PM 2.5, PM10, *NO*_2_, *SO*_2_ and *NO*_*x*_. This indicator is strongly correlated with COVID- 19 infections and death doubling times, where higher AQI tends to speed up case and death reporting. This result is consistent with other observational studies [57, 58]. Some authors propose that air pollution increases infectivity, as SARS-CoV-2 binds with airborne particulate matter [59–61] allowing the disease to persist for longer in the air. Although this should not be ruled out, as mentioned, air quality indicators also tend to proxy poor pulmonary health, which may increase death and case reporting, that is people with lung problems induced by air pollution are more likely to have symptomatic infections. It is well documented that long term exposure to certain pollutants has knock-on effects for people suffering from pulmonary viral infections [62–65]. For example, a study by Soukup et al. [66] found that regulated inflammatory responses to viral infections are altered by exposure to PM10, potentially increasing the spread of infection and therefore increasing viral pneumonia-related hospital admissions.

In general, case and death reporting doubling times were negatively associated with temperature. There is increasing evidence that COVID-19 is a seasonal disease [67, 68], especially in temperate climates where there are distinct seasonal phases i.e. summer and winter, with distinct temperature ranges, distinct levels of ultraviolet radiation (UV) and seasonal differences in air moisture carrying capacity. Although, it is important not to rule out physical factors influencing transmission, especially for long-distance transmission, given the nature of the disease (transmission mainly takes place over short distances in closed spaces), the influence of weather on human behavior is likely one of the major drivers of SARS-CoV-2 transmission. Weather is widely considered to influence people’s behavior [69] but research on this topic is surprisingly scant. According to Daniel et al., 2014 [70], people living in warmer / hotter locations, or during periods of warmer weather are more likely to employ a range of adaptive behaviors in response to warm and hot conditions i.e., keeping windows and doors open, use of wall and ceiling fans, air conditioning, which in turn may initiate a range of self-protective behaviors against SARS-CoV-2 transmission. Furthermore, warmer weather is also associated with recreational time spent outdoors [71] where SARS-CoV-2 transmission risk is likely to be lower. Although temperature also exhibited similar patterns for the death data model, it was only weakly statistically significant.

### 4.3 Limitations

Some of the limitations of the study are as follows. since the study relied on macrolevel data-driven approaches, it was limited to using aggregated data at the county level. With any analysis dealing with population-level areal data, issues can arise due to individual heterogeneity, which may lead to confounding bias, and it is not possible to draw causal inference due to the likely presence of endogeneity bias (e.g., omitted variable bias or reverse causality). Therefore, results were carefully evaluated from individual-level and clinical-based studies to draw conclusions. The use of further explanatory variables would have surely improved the study i.e. on homelessness, availability of Intensive Care Units (ICU), quality of medical facilities, and ratio of medical staff per person, but these data were not available. It is also important to note that given the unprecedented nature and scale of COVID- 19 outbreaks, data quality issues arise owing to the under-reporting of cases i.e., through under-diagnosis, lack of diagnostic tests and a lack of resources/time to carry out and implement mass testing. If data collection methods remained constant across counties over the time frame of this study, the calculation of doubling times can be a reliable measure. However, doubling times can be inflated by improving testing procedures i.e., better detection and reporting through the availability of better diagnostic tests, better sampling techniques, resource allocation, and increased awareness of the disease.

## 5 Conclusions

This paper investigated drivers of epidemic growth during the first wave of outbreaks in US counties, by assessing the association between COVID-19 epidemic doubling times with socio-economic, demographic, environmental factors, and government containment measures. Results suggest that the main drivers of new infections are higher population density, home overcrowding, manufacturing and recreation industries and poverty. By contrast, warmer temperatures slowed epidemic growth which was likely to be the result of human behavior responses to temperature. The main factors associated with death doubling times were age, poverty, air pollution and diabetes prevalence. Such findings help underpin current understanding of the disease epidemiology and also support current policy and advice recommending ventilation of homes, work-spaces and schools, along with social distancing and mask-wearing.

The results also suggest that states which adopted more stringent containment measures early on, did have some success suppressing the virus. We can presume that if this was replicated at a federal level, we would have seen much better outcomes in the United States. There are also numerous reports that there were huge failures at local level i.e. in care homes and business owners failing to protect residents and staff, by acting too late or not at all [72–74]. The results also show that those counties with the highest percentages of people with certain underlying health conditions, age, and poverty were also those which had higher death doubling times. Protecting these groups early on with income support schemes would have allowed the working vulnerable to stay at home and avoid infection [75, 76]. Furthermore, home overcrowding was also a very important factor in case doubling times and a policy of providing a quarantine location for those infected with SAR-CoV-2 would have surely slowed epidemic growth [77].

Finally, while it is not clear where the next threat will come from, anthropogenic activity like deforestation, wildlife trade, and intensive animal rearing, that encourages spillover from wild reservoirs, and influences the emergence and evolution of novel coronaviruses [9, 78, 79] will continue to present risks globally until better controls and regulations can be implemented [80]. If new coronaviruses emerge, with similar modes of transmission, we would hope that in future governments would quickly apply top-down measures to suppress the virus before more sophisticated measures can be implemented i.e. rapid community testing to isolate the infected. We hope this work will contribute to the scholarly debate and can shed light on some of the environmental and socio-economic factors driving SAR-COV-2 transmission.

## Data Availability

Embargoed until publiclation

## 5.1 Abbreviations

GDP: Gross Domestic Product
US: United States of America

## 5.2 Ethics approval and consent to participate

Not applicable

## 5.4 Data and code availability

An R project containing all the data and code that support the findings of this study is available in .Rdata format from https://doi.org/10.5281/zenodo.4994110.

## 5.5 Acknowledgments

Thanks to Panagiota Kotsila, Graham Mortyn, Victor Sarto Monteys and Cesira Urzi Brancati who provided advice and feedback on the research. Panagiota Kotsila, Graham Mortyn and Victor Sarto Monteys also supervised the project and are responsible for formulating ideas for the umbrella project Impacts of Climate Change (CC) on Human Health (HH) at ICTA-UAB: Integrating socio-economic and policy studies with natural science studies to enhance consilience of climate policy science.

This research was funded by ICTA’s Maria de Maeztu Unit of Excellence, awarded by the Spanish Ministry of Economy and Competitiveness. The award is the highest institutional recognition of scientific research in Spain.Thanks also to Patrizia Ziveri and Pedro Manuel Gonzalez Hernandez for supporting the project.

