## Additional file 2 for "Macro-Level Drivers of SARS-CoV-2 Transmission: A Data-Driven Analysis of Factors Contributing to Epidemic Growth During the First Wave of outbreaks in the United States"

### Supplementary Information

Matthew J. Watts

11/04/2020

#### Diagnostics: Infection model

S

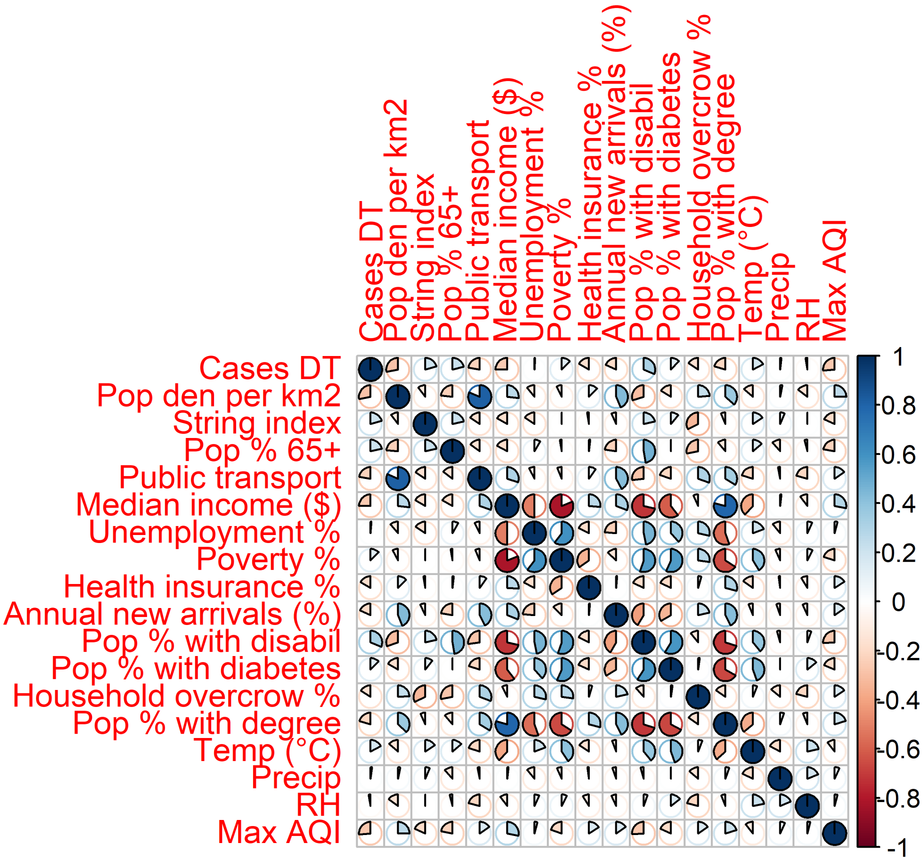


Figure S1: Correlation plot - infection model data.

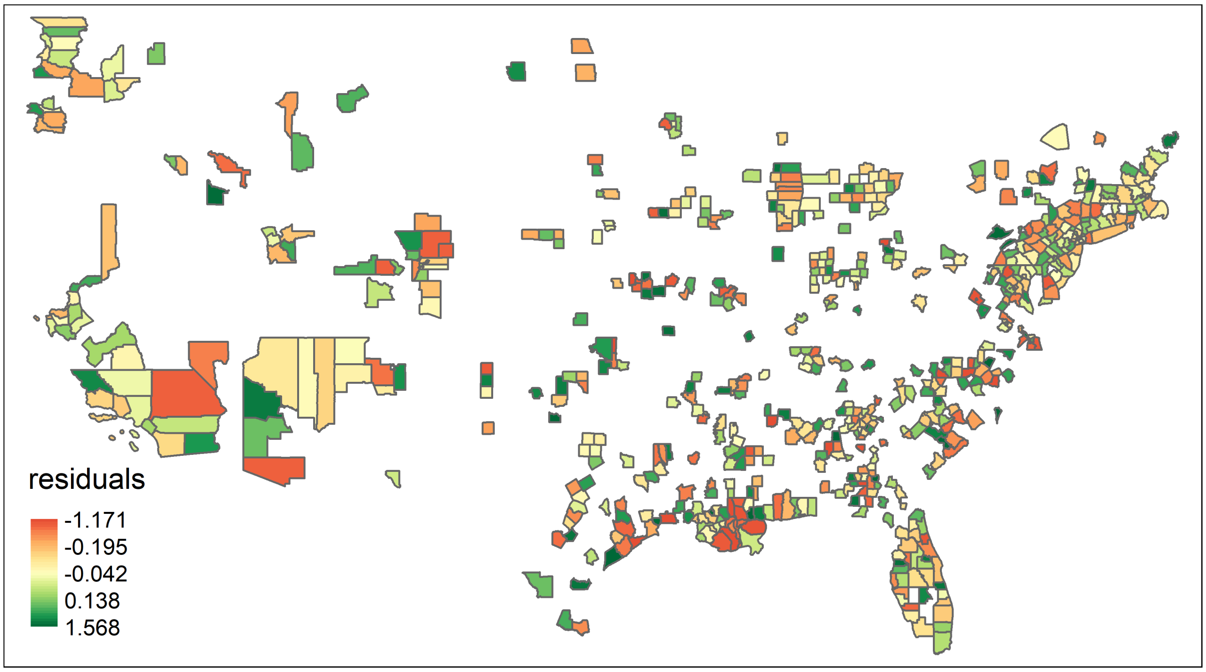


Figure S2: Spatial residuals - cases model.

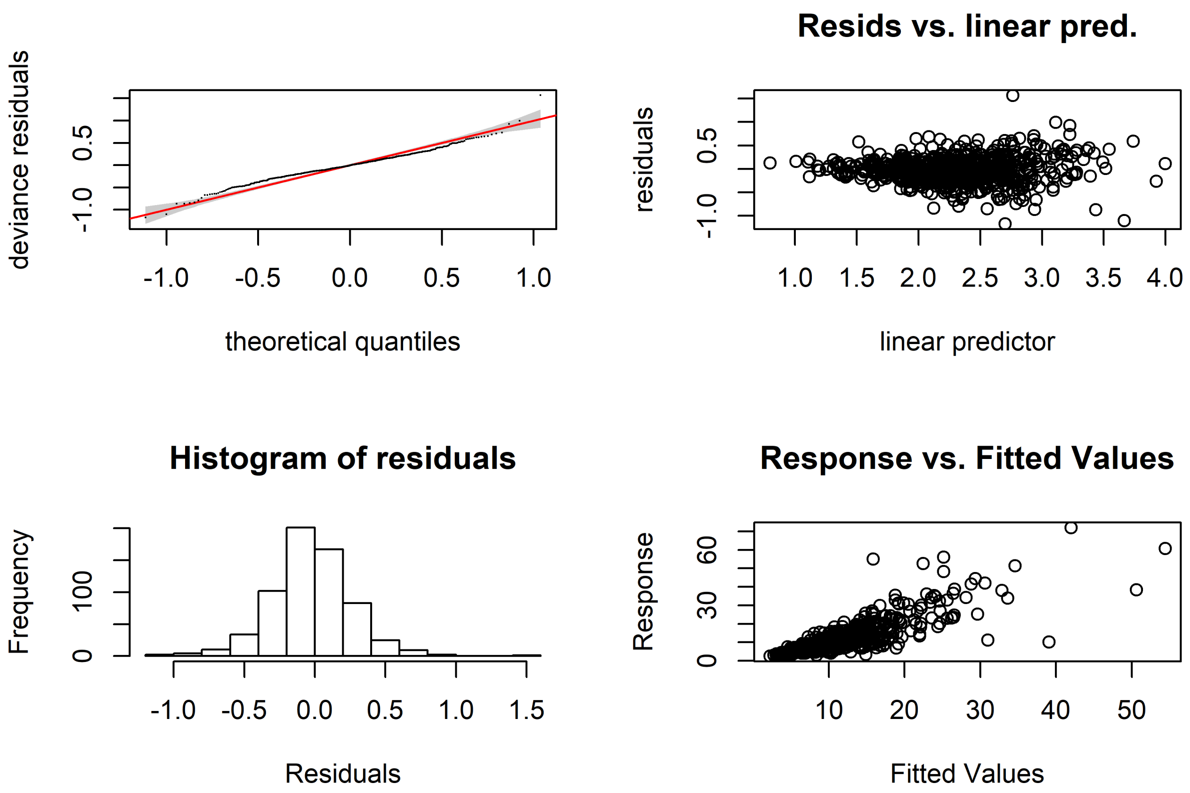


Figure S3: Model diagnostics - cases model.

#### Diagnostics: Mortality model

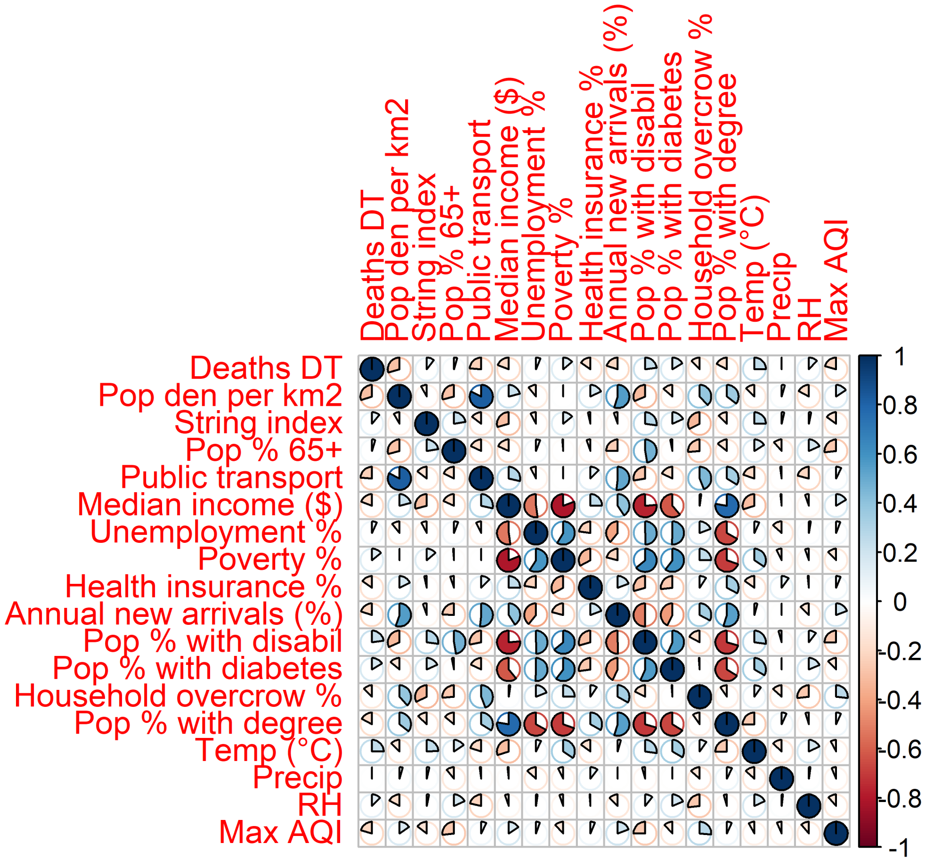


Figure S4: Correlation plot - mortality model data.

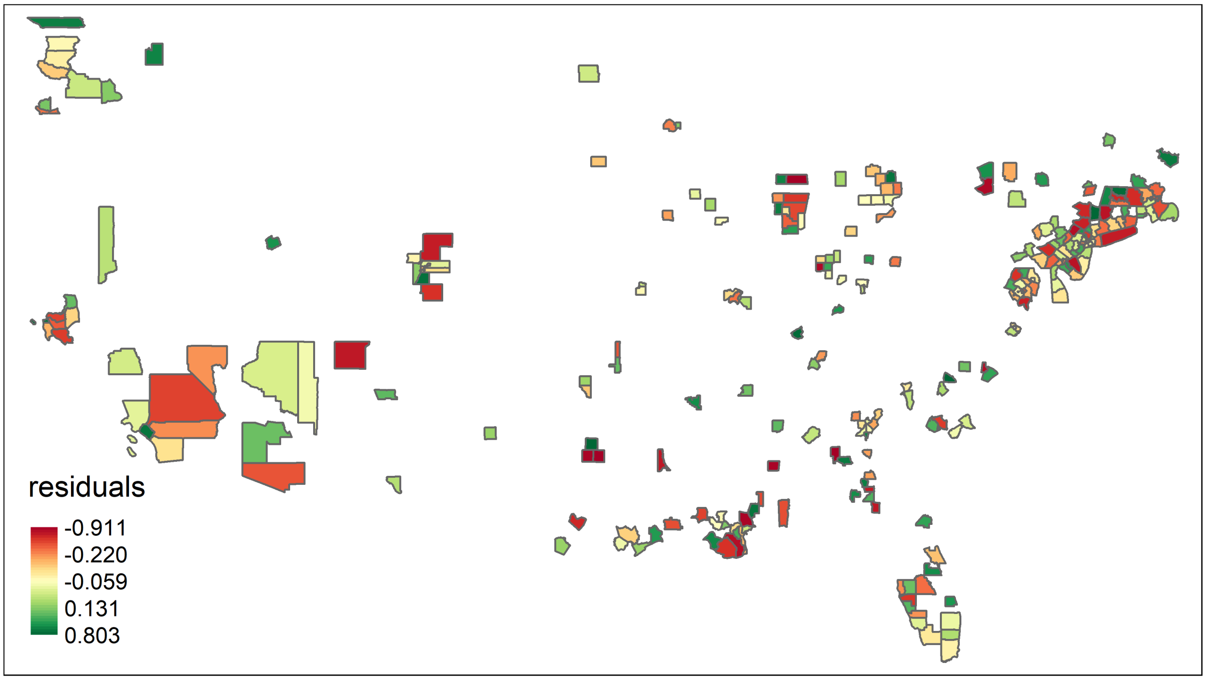


Figure S5: Spatial residuals - deaths model.

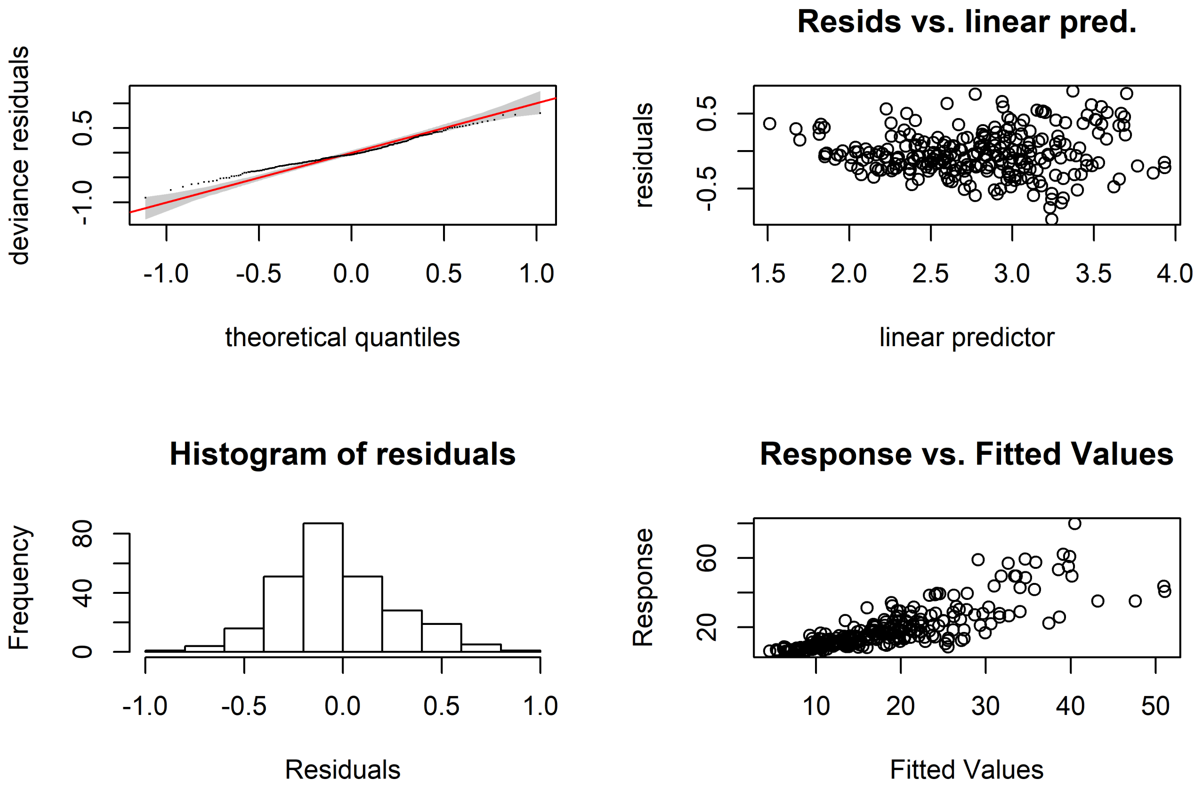


Figure S6: Model diagnostics - deaths model.
